# A Caveat to Using Wearable Sensor Data for COVID-19 Detection: The Role of Behavioral Change after Receipt of Test Results

**DOI:** 10.1101/2021.04.17.21255513

**Authors:** Jennifer L. Cleary, Yu Fang, Srijan Sen, Zhenke Wu

**Author notes:** **Corresponding author** Zhenke Wu, PhD. Department of Biostatistics, University of Michigan. 1415 Washington Heights, Ann Arbor, MI, 48109-2029, USA. These authors contributed equally: Jennifer L. Cleary, Yu Fang.

## Abstract

Recent studies indicate that wearable sensors have the potential to capture subtle within-person changes that signal SARS-CoV-2 infection. However, it remains unclear the extent to which observed discriminative performance is attributable to behavioral change after receiving test results. We conducted a retrospective study in a sample of medical interns who received COVID-19 test results from March to December 2020. Our data confirmed that sensor data were able to differentiate between symptomatic COVID-19 positive and negative individuals with good accuracy (area under the curve (AUC) = 0.75). However, removing post-result data substantially reduced discriminative capacity (0.75 to 0.63; delta= −0.12, p=0.013). Removing data in the symptomatic period prior to receipt of test results did not produce similar reductions in discriminative capacity. These findings suggest a meaningful proportion of the discriminative capacity of wearable sensor data for SARS-CoV-2 infection may be due to behavior change after receiving test results.

## Main

Recent studies ^1–4^ suggest enormous public health potential of wearable sensors in capturing subtle within-person changes that indicate an infection, such as by SARS-CoV-2. Detection of infection via wearable data provides a potentially effective, scalable method of infection surveillance, through passive, non-invasive methods ^5^. However, the assessments of wearable sensors for SARS-CoV-2 infection to date conflate two distinct streams of information - direct physiologic effects of infection and behavioral changes secondary to learning confirmation of infection through receipt of test results. Understanding the relative importance of these two streams of information in infection detection is critical to determining if infection surveillance may be possible through wearable technology. This paper seeks to further this understanding by leveraging a unique data set with individual-level dates of receiving test results that are linked to wearable data collected from a cohort of symptomatic COVID-19 positive or negative medical interns.

The Intern Health Study is a prospective cohort study that assesses mental health during the first year of residency training ^6,7^. Individuals starting residency in the 2019 and 2020 cohorts were invited to take part. Participating interns received a Fitbit Inspire HR or Charge 3 device (or $50 if they already have a Fitbit, Fitbit Inc., San Francisco, CA; or an Apple Watch, Apple Inc., Cupertino, CA) and $60 in compensation. The institutional review board at the University of Michigan approved the study. From April to December 2020, participants were sent multiple surveys that assessed whether they 1) exhibited any symptoms consistent with COVID-19 (e.g. fever, cough, shortness of breath, headache); (2) were tested for SARS-CoV-2 infection; (3) tested positive. Daily sleep duration, physical activity, and resting heart rate (RHR) were measured through Fitbit or Apple Watch throughout the first internship year. We focused on interns because this is a population that is likely to receive tests, receive test results quickly, and are more adherent to quarantine measures.

A total of 3,532 subjects participated in the 2019 and 2020 cohorts of Intern Health Study. Among them, 506 subjects experienced COVID-19-like symptoms between March 15 and December 2020 and of these, 379 reported being tested for SARS-CoV-2. There were 94 individuals who tested positive (“cases”) and 285 individuals who tested negative (“controls”). We included in the analysis 22 cases and 83 controls who had step, sleep, and RHR data available for more than 50% of the days during baseline (21 to 7 days prior to symptom onset) and test (0-7 days after symptom onset) periods, respectively (Extended Data Figure 1). Participants were on average 28.5 +/− 2.81 years of age, and 50.5% (n = 53) of the sample were female.

Our results are consistent with those reported by Quer et al. (2020) ^2^, validating the value of passive wearable sensor data in differentiating symptomatic COVID-19 positive from negative individuals. In particular, we followed Quer et al. (2020) ^2^ and used externally-constructed metrics that operationalize within-person changes in RHR, sleep, steps and all three combined. The metrics effectively contrast an individual’s wearable sensor data from the test period with those from the baseline period, which are then used to discriminate cases and controls. Using all the data in baseline and test periods (Figure 1a), we observed that metrics of within-individual change discriminated cases from controls except for RHR (Figure 2, a-d). Sleep minutes increased more among cases than controls after symptom onset (mean change: 47.9 in cases, 16.6 in controls p=0.044; area under the curve, AUC, based on SLEEPmetric = 0.66, 95% confidence interval, CI = 0.51-0.80). Cases reduced physical activity more than controls after symptom onset (mean change: −3,703 in cases, −1,038 in controls, p=0.002; AUC based on STEPmetric = 0.75, 95% CI = 0.63-0.87). Mean change in RHR is higher in the cases (1.3 in cases, 0.4 in controls, p=0.18) with the lowest discriminative ability based on RHRmetric (AUC = 0.63, 95% CI = 0.48-0.79). The combined metric based on all wearable sensor data results in an AUC of 0.75 (95% CI = 0.62-0.89).

**Figure 1.**
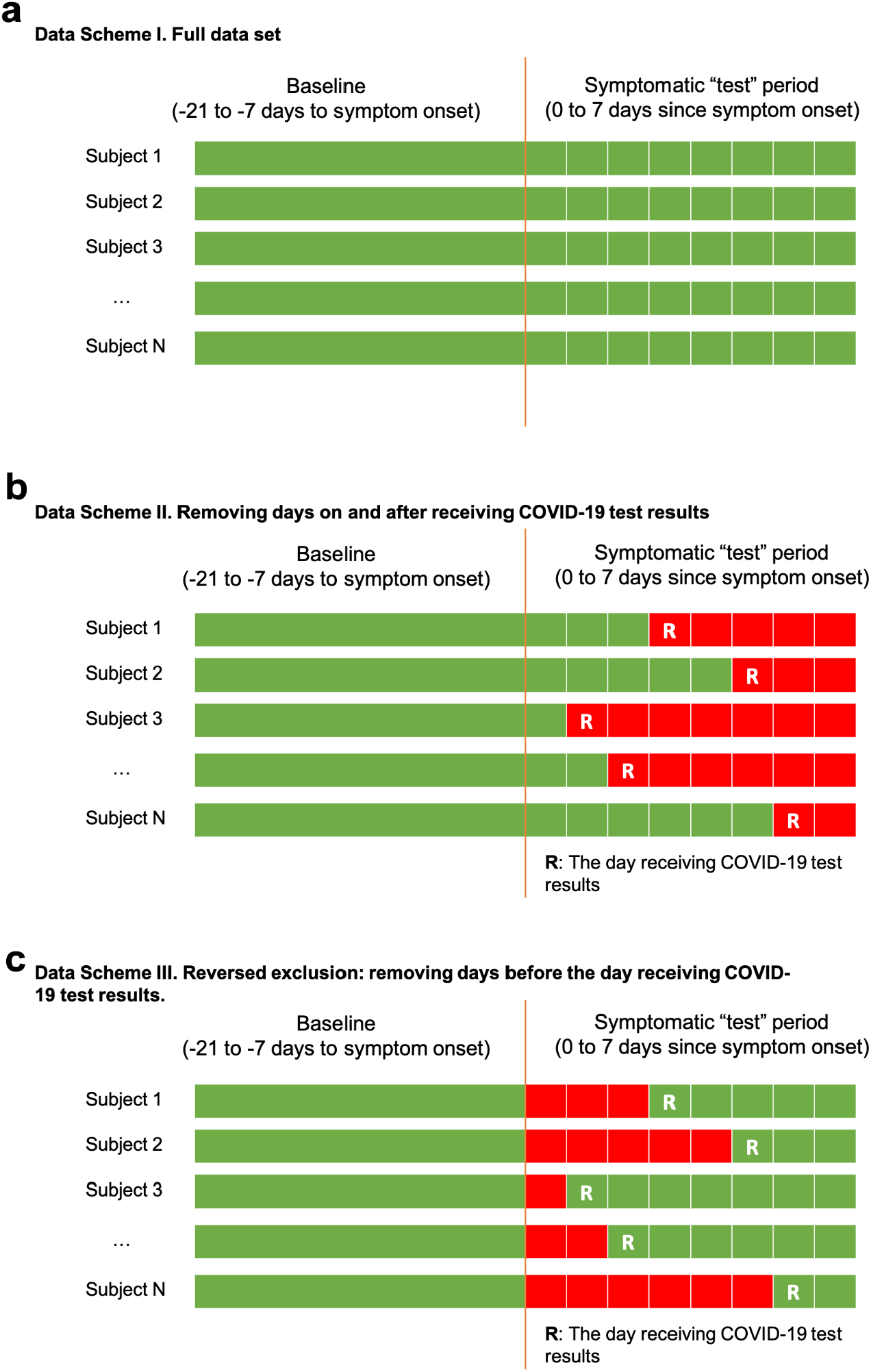
Data Schemes. **a-c**, green: included data; red: excluded data; R: the day receiving test results: (**a**) include all data; (**b**) exclude data on and after the day receiving test results; (**c**) exclude data before the day receiving test results since symptom onset. Ninety-two subjects (87.6%) received their results within the symptomatic period (0 to 7 days after symptom onset).

**Figure 2.**
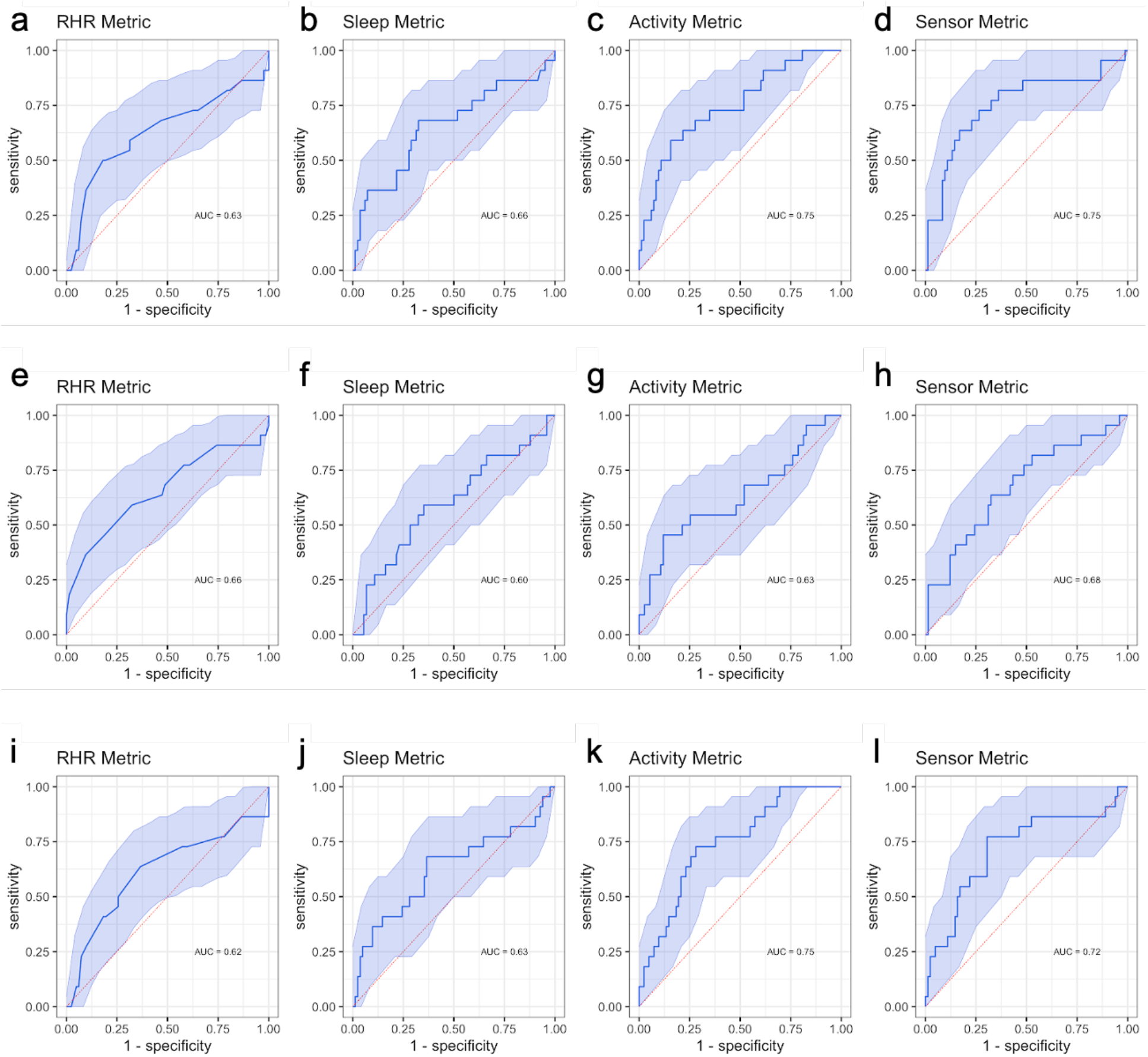
AUCs based on RHR, sleep, activity and all-sensor metric derived from wearable sensors to differentiate symptomatic subjects who were tested positive and negative corresponding to data schemes I-III. (**a-d**): Scheme I - all data; (**e-h**) Scheme II - remove data on and after knowing the test results; (**i-l**): Scheme III - remove data since symptom onset and before the test results. For each data scheme in the row, the four panels are for RHR, sleep, activity and sensor metrics, respectively.

To test whether the realized AUCs were mainly driven by the subset of data after receipt of test results, we conducted an analysis that removed data points on and after the result delivery date (Figure 1b). Compared with the previous analysis, we observed worse discriminative ability (Figure 2e-2h) by SLEEPmetric (AUC = 0.60, 95% CI = 0.42-0.76), STEPmetric (AUC = 0.63, 95% CI = 0.49-0.78), and combined sensor metrics (AUC = 0.68, 95% CI = 0.50-0.82), but similar performance in RHR (AUC = 0.66, 95% CI = 0.51-0.86). The AUC based on STEPmetric experienced the largest decrease (delta = −0.12).

To assess whether the observed decrease in AUC is consistent with random data removal or systematic information loss, we further conducted one-sided conditional permutation tests for each metric (see Methods). In particular, the test assesses the null that, compared to random data removal, no additional decrease in AUC is caused by systematically removing data after receipt of test results. For the STEPmetric, the observed decrease in AUC (step 2, Methods) stands in the left tail of the reference distribution of change in AUC (step 3, Methods; observed change in AUC: −0.12, p=0.013; Extended Data Figure 2c), indicating the observed decrease in AUC by removing post-result data is unlikely a chance event from data reduction and hence the importance of post-result data. Although cases and controls reduced average daily step counts after they became symptomatic, the reduction was significantly more among the cases after receipt of test results (mean change: cases −4,012, controls −1,016; p=0.001) and more so than during the symptomatic period before receipt of test results (mean change: cases −2,894, controls −1,083, p=0.03). For RHR and sleep metrics, we did not observe a statistically significant decrease in the AUC after removing the post-result data.

Finally, when only using the post-result-data in the test period (Figure 1c), the AUCs is comparable to the all-data AUC for all metrics (Figure 2i-2l, RHRmetric: 0.62 vs 0.63; SLEEPmetric: 0.63 vs 0.66; STEPmetric: 0.75 vs 0.75; all-sensor: 0.72 vs 0.75), indicating no substantial loss of discriminative accuracy is incurred by only using post-result data when defining the metrics. We performed conditional permutation tests as above, but with the number of random days removed being the number of days prior to receipt of test results. No statistically significant decrease in AUCs was observed for any of the metrics (Extended Data Figure 2e-2h).

Our analysis reveals the discriminative accuracy of wearable data in COVID-19 detection can be explained by behavior changes after receiving test results, more so driven by subjects’ within-person change in physical activity, less so by sleep or RHR. In particular, when removing data on and after receipt of test results, the AUC based on STEPmetric drops significantly from the all-data AUC. A small though non-statistically significant drop was observed for SLEEPmetric. No decrease was observed for physiology-based RHRmetric. This pattern is consistent with behavior change after receiving COVID-19 test results. Compared to symptomatic individuals who tested negative, symptomatic individuals who received a positive COVID-19 test may initiate stricter quarantine measures thus reducing activity and aim to get more sleep. It appears that in the short term sleep is more resistant to change than physical activity during the test period, likely strongly regulated by circadian rhythms ^8^.

This study has some limitations. First, our sample is a small subset of symptomatic subjects from a sizable cohort. In future studies, it is critical to aggregate data from multiple studies to further validate and study the variation in AUC with factors that may impact the propensity of behavioral change. Second, the cohort is likely not representative of the entire spectrum of population that may have access to both wearables and tests. However, the unique cohort of medical interns who are likely more adherent to quarantine measures strengthened the specific investigation addressed here. It is of interest to investigate the same question in a broader population. Third, the SARS-CoV-2 tests are not perfectly sensitive or specific. Knowledge about these test-related parameters will likely further improve AUC estimates. Fourth, recall of symptom onset date and test date might not be entirely accurate, but this population of medical interns is particularly primed to remember the dates due to workplace enforcements of symptom screening, testing, and compulsory quarantines.

In a future pandemic, passively-collected wearable data linked with test results may reveal distinct patterns of behavioral change across subpopulations. For example, lack of appropriate behavioral changes upon receiving test results may hurt discriminative accuracy based on wearable sensor data. Variation in the AUC of the step metric by age group may indicate differential levels of within-person change in activity. Groups with higher step-based AUC may have effectively quarantined after receiving their test results; while groups with lower step-based AUC may indicate either delay in their receiving the test results or difficulty and infeasibility in reducing physical activity. Subpopulations with lower observed AUCs may benefit from more targeted public health policy innovations that may promote behavioral change, such as self-quarantine measures.

## Data Availability

Data for replication the results in the paper is available upon reasonable request.

## Acknowledgements

This study was supported by grants from the National Institute of Mental Health (R01 MH101459) to S.S. and Z.W. and an investigator grant from Precision Health Initiative at University of Michigan, Ann Arbor to Z.W. and S.S.. J.C. was supported by T32HD007109. We thank the interns and residency programs who took part in this study.

## Author Contributions

J.C., Y.F., S.S., Z.W. made substantial contributions to the study conception and design. S.S. made substantial contributions to the acquisition of data. J.C., Y.F., Z.W. conducted statistical analysis. J.C., Y.F., S.S., Z.W. made substantial contributions to the interpretation of data. J.C., Y.F., Z.W. drafted the first version of the manuscript. J.C., Y.F., S.S., Z.W. contributed to critical revisions and approved the final version of the manuscript. J.C., Y.F., S.S., Z.W. take responsibility for the integrity of the work.

## Competing Interests Statement

The authors have no competing interests to disclose.

## Data and code availability

Deidentified data supporting the results and figures in this manuscript are available upon reasonable request and completion of a data agreement with the Intern Health Study team. Code for data preprocessing and statistical analysis is available upon request.

## Methods

### Metrics Definition

Participants were drawn from the 2019 and 2020 cohorts of the Intern Health Study. Study recruitment and procedures are detailed elsewhere.^8^ Briefly, incoming first-year medical residents were surveyed throughout the pandemic from April to December 2020 and asked to report whether and when they experienced any potential COVID-19 symptoms, were tested, and their test results. The sample for this analysis included individuals who reported symptoms and a COVID-19 test, as well as at least 50% of the wearable data (collected through Fitbit or Apple Watch) during both baseline (21 to 7 days prior to symptom onset) and test (0 to 7 days after symptom onset) periods.

Following Quer et al. (2020) ^2^, we calculated metrics for sleep, activity, and resting heart rate (RHR), as well as an overall wearable sensor metric for each participant:

**RHRmetric** = max(dailyRHR[test]) − median(dailyRHR[baseline])/IQR

**SLEEPmetric** = mean(dailySLEEP[test]) − median(dailySLEEP[baseline])/IQR

**STEPmetric** = mean(dailySTEP[test]) − median(dailySTEP[baseline])/IQR

**SENSORmetric** = RHRmetric /10 + SLEEPmetric − STEPmetric

### Discriminative Accuracy

We calculated ROC curves, AUC, sensitivity (SE), specificity (SP) for each metric to compare the intra-individual change in each metric with symptom onset between COVID-19 positive and COVID-19 negative individuals. To assess which part of the test period data is mainly responsible for the realized AUC, we calculated these parameters in three data schemes: Scheme I - using all the data in baseline and test periods; Scheme II - removing data on and after receipt of test results in test periods; Scheme III - removing data before receipt of test results in test periods.

### Conditional Permutation Tests

In order to test the statistical significance of the observed AUC decrease in Scheme II and III, we designed the one-sided conditional permutation tests in a way that breaks the link between the indices of days removed during the test period and the dates of receiving the test results hence creating a null distribution that is adequate for assessing the statistical significance of the observed change in AUC. In particular, for each metric (RHR, sleep, activity, sensor) we perform the following steps:

Step 1. Calculate AUC based on all the baseline and test data;

Step 2. Remove part of the test data (on/after receiving the test results as in Figure 1b; OR before receiving the test results as in Figure 1c), and calculate a single AUC and the change from the AUC in Step 1;

Step 3. Create B=1000 data sets, each by randomly removing the same amount of data for each person as in Step 2; based on each of B random reduced data sets, calculate an AUC and the difference from the AUC in Step 1, resulting in B=1000 values of change in AUC;

Step 4. Compare the change of AUC in Step 2 against the null distribution of the change of AUCs in Step 3; Calculate the p-value by the observed fraction among the 1000 randomly reduced data sets that have AUC change less than or equal to the observed change in Step 2.

All analyses were conducted using R 4.0.2 ^9^.

**Extended Data Figure 1.**
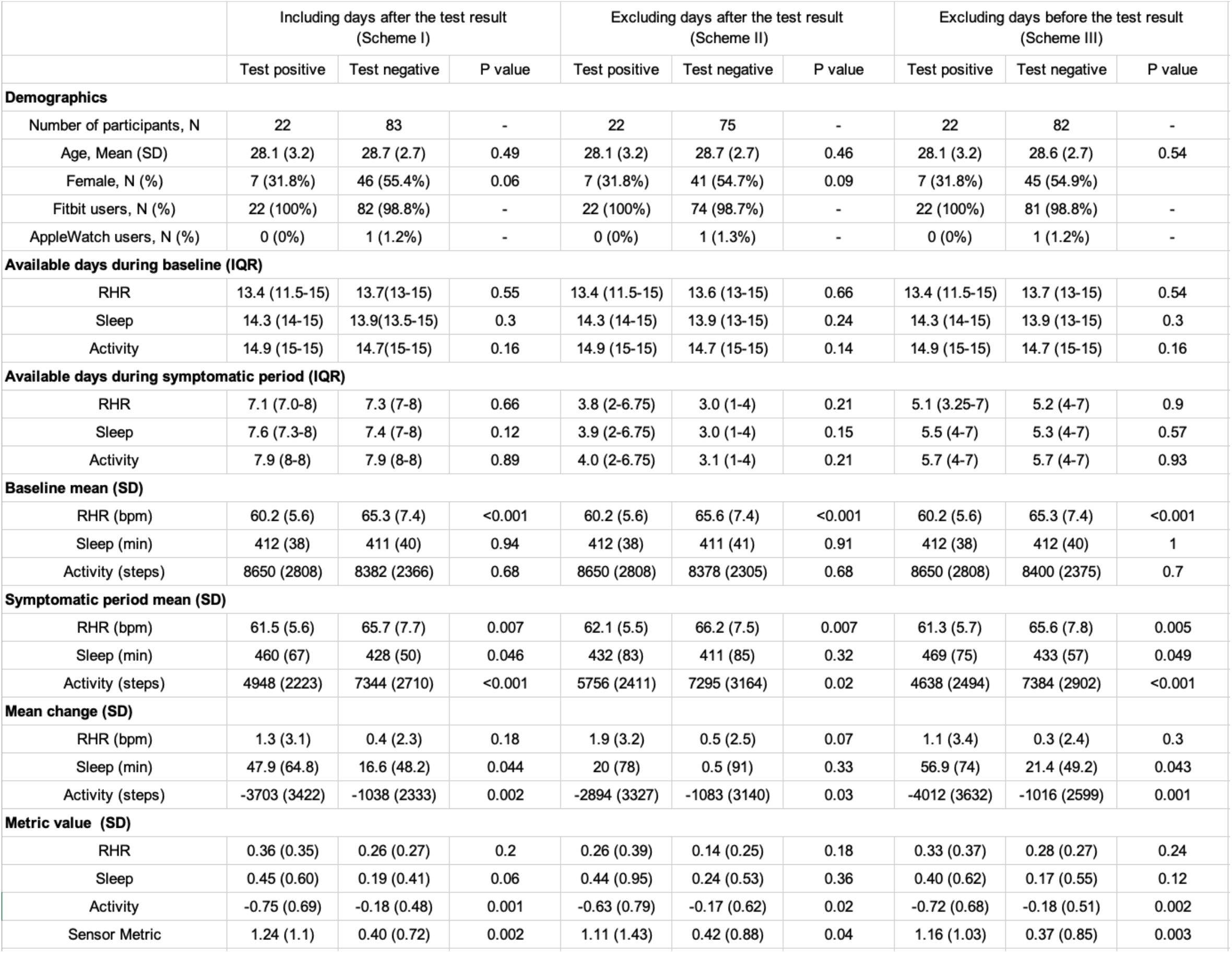
Summary of key characteristics, metrics and COVID-19 test results.

**Extended Data Figure 2.**
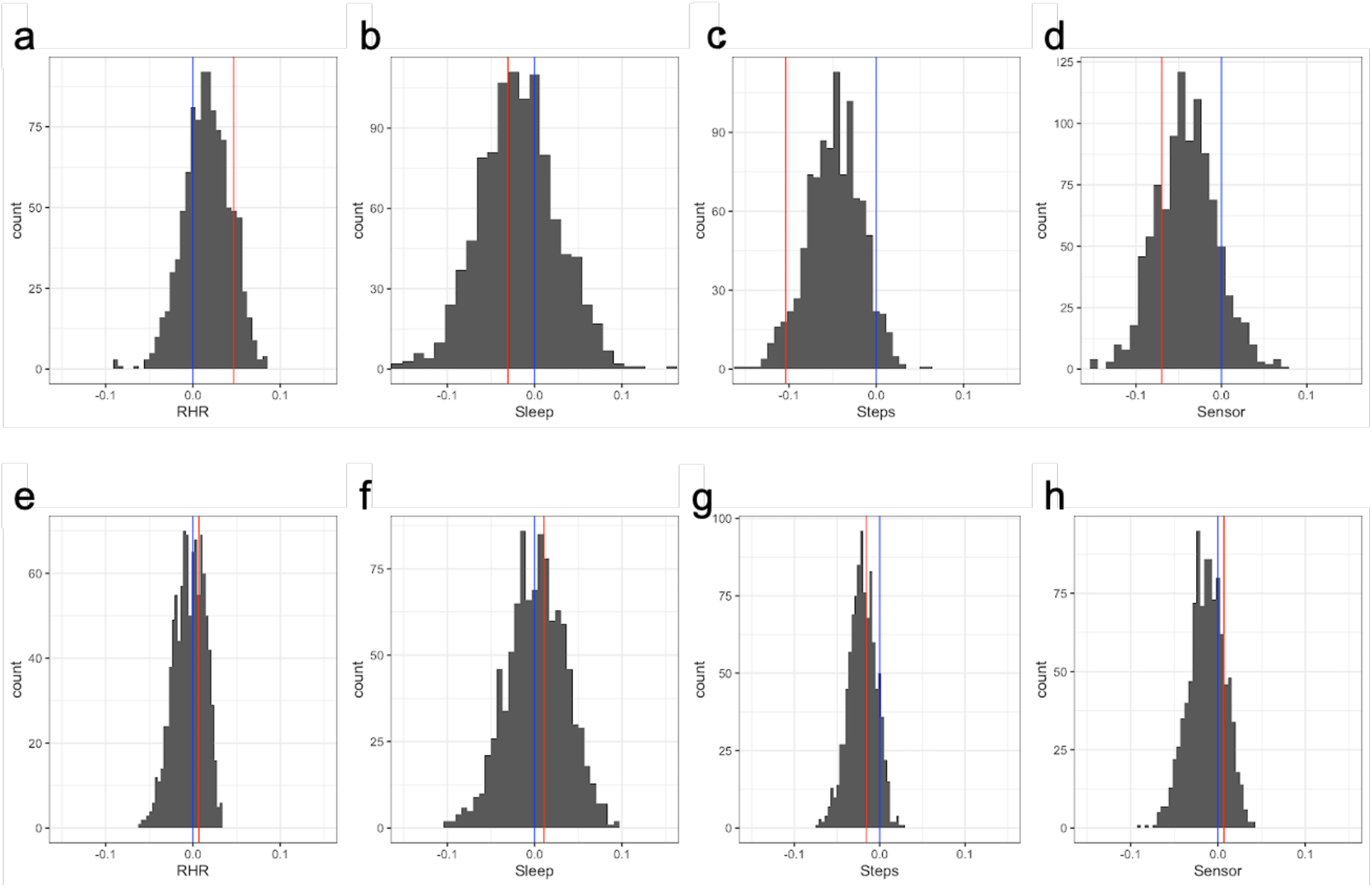
One-sided conditional permutation test for assessing the null that, compared to random data reduction, no additional change in AUC caused by removing data: (**a-d**) on or after receipt of test result and (**e-h**) in the symptomatic period and prior to receipt of test results. The random data removal and AUC calculation are done for RHR, sleep, step and the all-sensor data, respectively (shown in four panels in each row). In each panel, the red line indicates the observed change of AUC; the blue line is at zero, indicating no change. The reference distributions are not centered at zero despite data removal being random; because on average there are more days after the receipt of test results than before, random data removal may still impact AUCs. For each metric, if a red line is at the left tail of the histogram, we conclude a statistically significant additional decrease in AUC.

## Notes

### Competing Interest Statement

The authors have declared no competing interest.

### Author Declarations

IRB, University of Michigan, Ann Arbor

